# Developing a Comprehensive Framework for Real-World Data Case Validation in Vaccine Safety Monitoring: The VAC4EU experience

**DOI:** 10.1101/2025.08.25.25334384

**Authors:** Amirreza Dehghan Tarazjani, Daniel Weibel, Taylor Aurelius, Laura Zwiers, Jesse van den Berg, Lina Pérez-Breva, Antonio Gimeno-Miguel, Luca Stona, Martín Solórzano, Anteneh Assefa Desalegn, Beatriz Poblador-Plou, Thom Lysen, Jannik Wheler, Mahmoud Zidan, Juan José Carreras, Felipe Villalobos, Vera Ehrenstein, Kathryn Morton, Cristina Rebordosa, Fariba Ahmadizar, Joan Fortuny, Alejandro Arana, Miriam Sturkenboom

## Abstract

**Purpose:** Real-world evidence (RWE) is essential for post-licensure monitoring of vaccine safety, as pre-licensure trials are often limited in sample size, population diversity, and duration of follow-up. The Vaccine Monitoring Collaboration for Europe (VAC4EU) has developed a structured validation pipeline that operationalizes Brighton Collaboration (BC) case definitions for real-world data (RWD) in a harmonized, scalable and reusable manner.

**Methods:** VAC4EU developed a systematic, stepwise approach to validate vaccine safety outcomes utilizing BC case definitions where available. The approach involves: 1) Critical review of BC definition and adaptation to RWD by clinical and RWD experts; 2) Creation of dummy cases based on published reports; 3) Creation of REDCap electronic data collection forms (eDCF) incorporating decision logic to assigned levels of certainty (LOC); 4) Iterative testing of decision logic; and 5) comprehensive training of abstractors with real-time feedback. A dedicated task force assigned reference LOCs for dummy cases. Inter-rater reliability was measured using Fleiss’ kappa (κ) by comparing abstractor LOCs to the reference standard.

**Results:** The validation pipeline was applied to 16 COVID-19 vaccine safety outcomes, 13 of them had existing BC definitions. Eleven eDCFs required adaptation to accommodate missing or unstructured information common in RWD. In total, 78 dummy cases were developed across the 16 outcomes, and 15 REDCap eDCFs were created. Myocarditis and pericarditis shared a single form. Across 33 trained abstractors, 747 individual case abstractions were completed. Agreement analysis showed 93 discrepancies (12.4%) and moderate overall concordance (κ = 0.55), with the lowest for Thrombosis with Thrombocytopenia Syndrome (κ = –0.05).

**Conclusion:** The VAC4EU validation pipeline provides a standardized framework for training and validating vaccine safety outcome validation using RWD. By adapting BC case definitions and emphasizing targeted abstractor training with ongoing feedback, this approach improves the reliability of post-marketing surveillance studies.

## Introduction

Vaccines prevent millions of deaths and diseases annually ^1^. Monitoring their benefit-risk balance is essential throughout the product lifecycle ^2^. As vaccines are administered to generally healthy individuals, the tolerance for safety risks is much lower compared with therapeutic products In this context, real-world evidence (RWE) derived from real-world data (RWD) plays a crucial role in evaluating the safety and effectiveness of vaccines, especially in the post-licensure phase. Therefore, the validation of safety outcomes is a key element in post-marketing surveillance, ensuring that rare or unexpected adverse events are accurately identified and assessed.

Health authorities such as the European Medicines Agency (EMA) and the U.S. Food and Drug Administration (FDA) now widely support the use of real-world data (RWD) in vaccine safety studies, provided that outcomes are rigorously validated to ensure data reliability, relevance, and traceability ^3^. In particular, the FDA’s recent guidance on RWD and RWE underscores the critical role of validation processes in regulatory decision-making, particularly when using electronic health records (EHRs) and medical claims data ^3^. The guidance emphasizes the importance of data reliability, defined as the accuracy and consistency of data over time; relevance, referring to how well the data elements reflect the concepts of interest; and traceability, which allows verification of the data source and transformations. It prioritizes comprehensive data capture, mitigation of missing information, and effective data linkage. The guidance also recommends evaluating whether data sources adequately represent study populations, exposures, and outcomes. Suggested validation approaches include complete verification or performance assessments through sampling strategies, tailored to specific study contexts ^3^.

The European Network of Centers in Pharmacoepidemiology & Pharmacovigilance (ENCePP) Methods Guide highlights the potential impact of misclassification of exposure, outcomes and covariates on study results and advocates for estimating sensitivity, specificity, and positive predictive values ^4^. Such misclassification can introduce bias into effect estimates; for instance, differential misclassification may lead to either underestimation or overestimation of associations, thereby compromising internal validity ^5^. Dremains the gold standard for outcome validation, whereas algorithm-based or indirect approaches may be acceptable alternatives when chart review is not feasible ^6,7^.

The EMA’s Guideline on Good Pharmacovigilance Practices (GVP) emphasizes the use of routinely collected healthcare data, such as electronic health records, disease registries, hospital discharge databases, or insurance claims, for large-scale studies, despite challenges like incomplete data and limited follow-up. The EMA stresses selecting data sources that balance validity and efficiency while adhering to privacy regulations ^8^.

In the framework of vaccine safety regulation, the Brighton Collaboration (BC) case definitions are considered standard for harmonized vaccine safety assessment and are intended to be applicable in different types of data collections: prospectively in clinical trials, during passive reporting, and retrospectively in surveillance systems or RWD ^9^. However, applying BC case definitions retrospectively to RWD sources in heterogeneous data source settings is not always straightforward, particularly in multinational studies where differences in healthcare systems and abstractors’ interpretation of criteria can lead to inconsistencies in case validation, as was observed in, for example, the H1N1 pandemic vaccine safety studies on Guillain-Barré Syndrome (GBS) and narcolepsy ^10–12^.

The Vaccine Monitoring Collaboration for Europe (VAC4EU) is an ongoing initiative aimed at generating high-quality evidence on vaccines to support regulatory decision-making in worldwide. VAC4EU developed a harmonized process for validating outcomes across heterogeneous RWD sources, including preparing data and abstractors to conduct validation. This paper describes how VAC4EU adapted case definitions for RWD, and trained abstractors for outcome validation, and describes performance of case validation across VAC4EU studies for interrater variability and validation accuracy.

## Methods

### Setting

VAC4EU facilitates the implementation of vaccine studies with RWD in a distributed manner across different organizations (data expert and access providers, DEAPs). DEAPs used the ConcePTION common data model (CDM) to transform their data ^13^. Through this framework, study partners are able to collaborate across different types of RWD sources by sharing common protocols and analytical scripts, while the data are kept local. DEAPs processed the data under local implementation of the General Data Protection Regulation (GDPR) ^14^. A stepwise process was developed to implement validation of the identified potential cases through retrospective review of local source data in a harmonized and federated manner.

The step-by-step workflow of this pipeline is illustrated in Figure 1. Each validation process step is described below.

**Figure 1.**
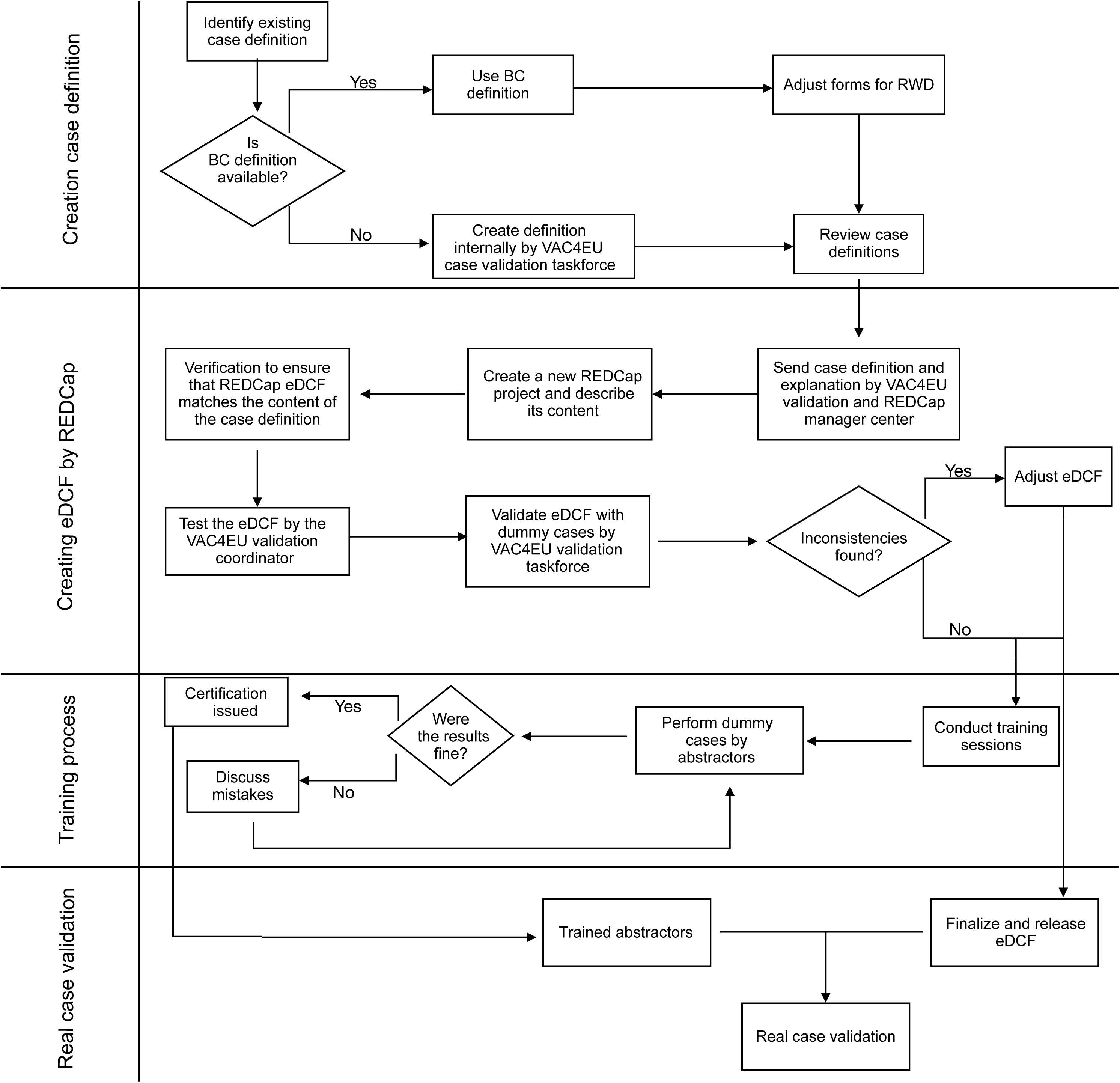
VAC4EU Case Validation Workflow.

#### 1. Definitions of Outcome Variables

The initial step in the validation process involves the identification of case definitions for outcomes. VAC4EU adopted the principle to use BC case definitions. If BC definitions were not available, the VAC4EU validation task force (comprised of principal investigators of studies and medically trained persons) created definitions based on literature review.

#### 2. Review and Adaptation of Data Extraction Forms

The VAC4EU validation task force conducted a comprehensive review of publicly available data extraction forms from the BC case definition companion guides and adapted them, where necessary, to better align with data likely to be available in a RWD setting. BC case definitions were originally developed for use in controlled clinical settings such as randomized controlled trials (RCTs), passive surveillance systems, and detailed case investigations, where comprehensive clinical, laboratory, and imaging information is often available. A key challenge addressed during this phase was to define how to handle the absence of information, which in RWD can mean actual absence of the condition or that the presence of the condition has not been recorded. In general, in RWD, there is information on which procedures, such as diagnostic tests, imaging, or treatments, were done and which diagnoses were set, but limited information on which conditions were actively excluded, or which procedures were not performed. This reality in RWD required the creation of guidelines for data abstraction to ensure consistent handling of the absence of information. The VAC4EU validation task force also aimed to make the data extraction questionnaires as straightforward as possible for abstractors. Simplified question formats were used to minimize complexity, and specific guidance documents were provided to assist abstractors.

#### 3. REDCap Data Collection Tool

VAC4EU uses the REDCap system for data collection to deploy a common data collection structure while keeping data local ^15,16^. The REDCap system was set up by Vall d’Hebron Hospital as the VAC4EU REDCap managing center during the development and testing phases. A review of the electronic data collection form (eDCF) was conducted by the VAC4EU validation task force and REDCap managing center, incorporating feedback from training sessions with dummy cases to refine the process and validate system functionality before full implementation.

#### 4. Dummy Cases

The VAC4EU validation task force created a set of five dummy case narratives for each outcome for training abstractors and assessment of the REDCap eDCFs. These cases were adapted from published case reports and modified to ensure blinding to vaccination status. Each case was designed to represent a different Level of Certainty (LOC), allowing full coverage of all LOC categories during training. The VAC4EU validation task force members completed the REDCap forms that were created by the REDCap managing center for each of the dummy cases to assess whether the eDCF captured the required information accurately. The dummy cases review was performed in a REDCap system hosted locally by each DEAP. The VAC4EU validation coordinator (AD) reviewed the results, comparing responses across task force members to identify any inconsistencies or issues. Discrepancies in the assigned LOCs were discussed. The consensus LOC for each case was determined through agreement among all task force members after discussion. Any fields or logic that caused confusion or produced varied responses were discussed, and necessary adjustments were made to improve clarity and accuracy.

#### 5. Training of Abstractors

The VAC4EU validation coordinator conducted training sessions to train validation abstractors from each DEAP on how to use the data collection forms and guidelines, discuss complex items, and resolve questions through examples. Training included:

- Introduction to the eDCF and data abstraction process.
- Practical exercises with 5 dummy cases for each outcome.
- Review of completed dummy cases and feedback by the VAC4EU validation coordinator.

Individual training certificates were issued after successfully completing the training. A follow-up meeting was held to discuss any doubts or challenges encountered during the dummy cases validation, particularly for cases where discrepancies were observed between the abstractors’ results and the expected outcomes defined by the VAC4EU validation task force.

#### 6. Finalization of eDCF

Based on insights gained during the training, the VAC4EU validation task force reviewed the abstractors’ feedback. Where necessary, adjustments were made to the eDCF or instructions to improve usability. After incorporating these refinements, the VAC4EU validation task force released the final eDCF for use in the study specific case validation.

#### 7. Support and Quality Control During study

During the study-specific case validation, the VAC4EU validation task force provided ongoing support to abstractors, addressing their questions and uncertainties as they arose. To maintain consistency and ensure alignment across DEAPs, the task force held weekly meetings to discuss common challenges and review emerging issues.

### Analysis

We assessed the agreement between abstractors (inter-rater reliability) across all outcomes using Fleiss’ Kappa ^17^, suitable in settings with more than two abstractors per outcome. For each outcome, a total of 4 to 22 abstractors were involved across all participating DEAPs combined.

The observed agreement (P_o_) was calculated for each case as the proportion of abstractors who selected the correct LOC classification. The overall observed agreement for each dataset was then computed as the average of the observed agreements across all cases.

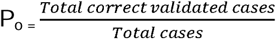

The expected agreement (P_e_) was calculated across all abstractors and cases as follows:

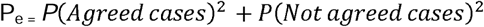

The Fleiss’ Kappa statistic was then computed using the following formula:

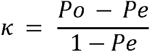

A higher Fleiss’ Kappa indicated stronger agreement between abstractors than expected by chance. Kappa values were categorized as ≤ 0 (no agreement), 0.01–0.20 (slight agreement), 0.21–0.40 as fair, 0.41– 0.60 as moderate, 0.61–0.80 as substantial, and 0.81–1.00 as almost high agreement ^18^. To estimate the precision of the Fleiss’ Kappa statistic, the standard error (SE) was calculated using the formula:

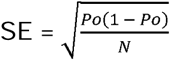

Where Po is the observed agreement and N is the number of cases in the dataset. The 95% confidence interval (CI) for Fleiss’ Kappa was then calculated by multiplying the standard error by the Z-score corresponding to a 95% confidence level (1.96) and adding this value from the Kappa estimate:

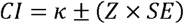

All calculations were performed using R (packages: irr).^19,20^

## Results

For five PASS on COVID-19 vaccines (EUPAS numbers 41623, 47708, 45362, 105009 and 43556), VAC4EU conducted case validation for GBS, narcolepsy, idiopathic Thrombocytopenic Purpura (ITP), thrombosis with thrombocytopenia syndrome (TTS), thrombocytopenia with bleeding, myocarditis, pericarditis, transverse myelitis, anaphylaxis, major congenital anomalies, encephalopathy (including acute disseminated encephalomyelitis (ADEM)), deep vein thrombosis (DVT), pulmonary embolism (PE), hemorrhagic ischemia, non-hemorrhagic ischemia, and cerebral venous sinus thrombosis (CVST).

For GBS, narcolepsy, ITP, TTS, myocarditis, pericarditis, transverse myelitis, anaphylaxis, major congenital anomalies, encephalopathy (including ADEM), DVT, pulmonary embolism, and CVST, a BC definition was available. For the remaining 3 outcomes (hemorrhagic stroke, non-hemorrhagic stroke, and thrombocytopenia with bleeding), data collection forms were developed based on literature review. Table 1 summarizes how BC definitions were adapted for use in the VAC4EU eDCFs. For each outcome, the total number of criteria implemented reflects both unmodified BC criteria and those either newly introduced or adapted to suit RWD settings. The “Adapted BC Criteria” column indicates how many original BC items required changes for clarity, relevance, or feasibility in RWD abstraction, while the “New Criteria” column captures additional items created by the task force. One such example is the criterion “Outcome reported by a specialist, but without accompanying clinical details”, which was added to capture situations where a diagnosis is stated but lacks sufficient clinical documentation to fully assess the case. Most importantly, the task force distinguished between diagnoses made by a specialist without further details and cases where there was insufficient information to meet the case definition. This was achieved by adding a subgroup to level 4, labeling it as 4a, and classifying cases with insufficient information to meet the case definition as 4b.

**Table 1.**
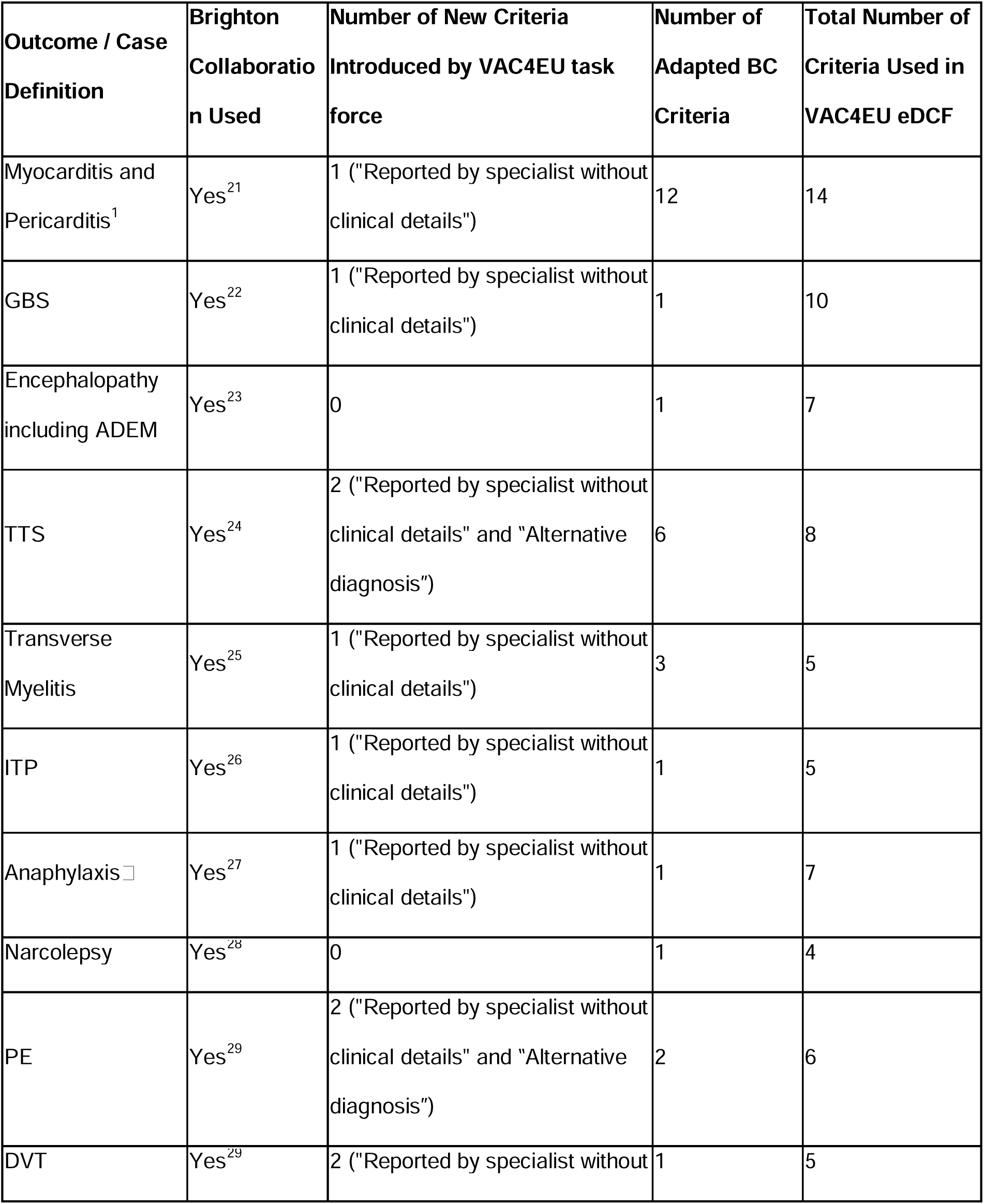
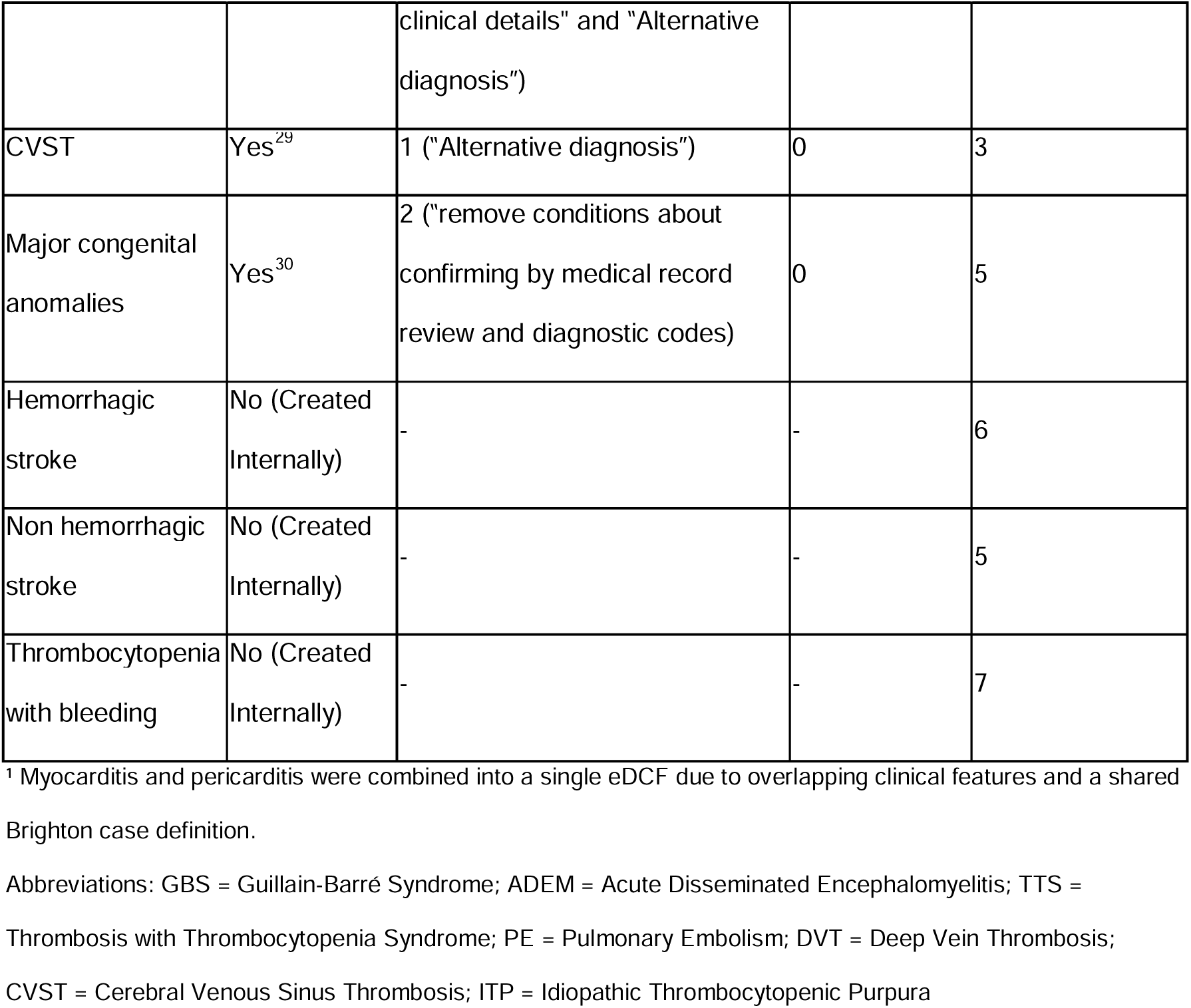
Adaptations to Case Definitions, Introduction of New Criteria by the VAC4EU Validation Task Force.

For the 16 outcomes, 78 dummy case narratives were created. In total, 33 abstractors were trained. The distribution of abstractors across different outcomes is summarized in Table 2. In total, 747 dummy cases were classified by these 33 abstractors and reviewed by the VAC4EU validation coordinator. Most abstractors were trained only on a subset of outcomes relevant to their assigned outcomes. As a result, each abstractor completed only the dummy cases corresponding to the specific outcome(s) they would validate. Discrepancies between the abstractors’ results and the expected outcomes defined by the VAC4EU task force were observed in 93 dummy cases (12.40%). The Fleiss’ Kappa values for the level of agreement between abstractors, across all outcomes, are reported in Table 2, along with their corresponding 95% CI.

**Table 2.**
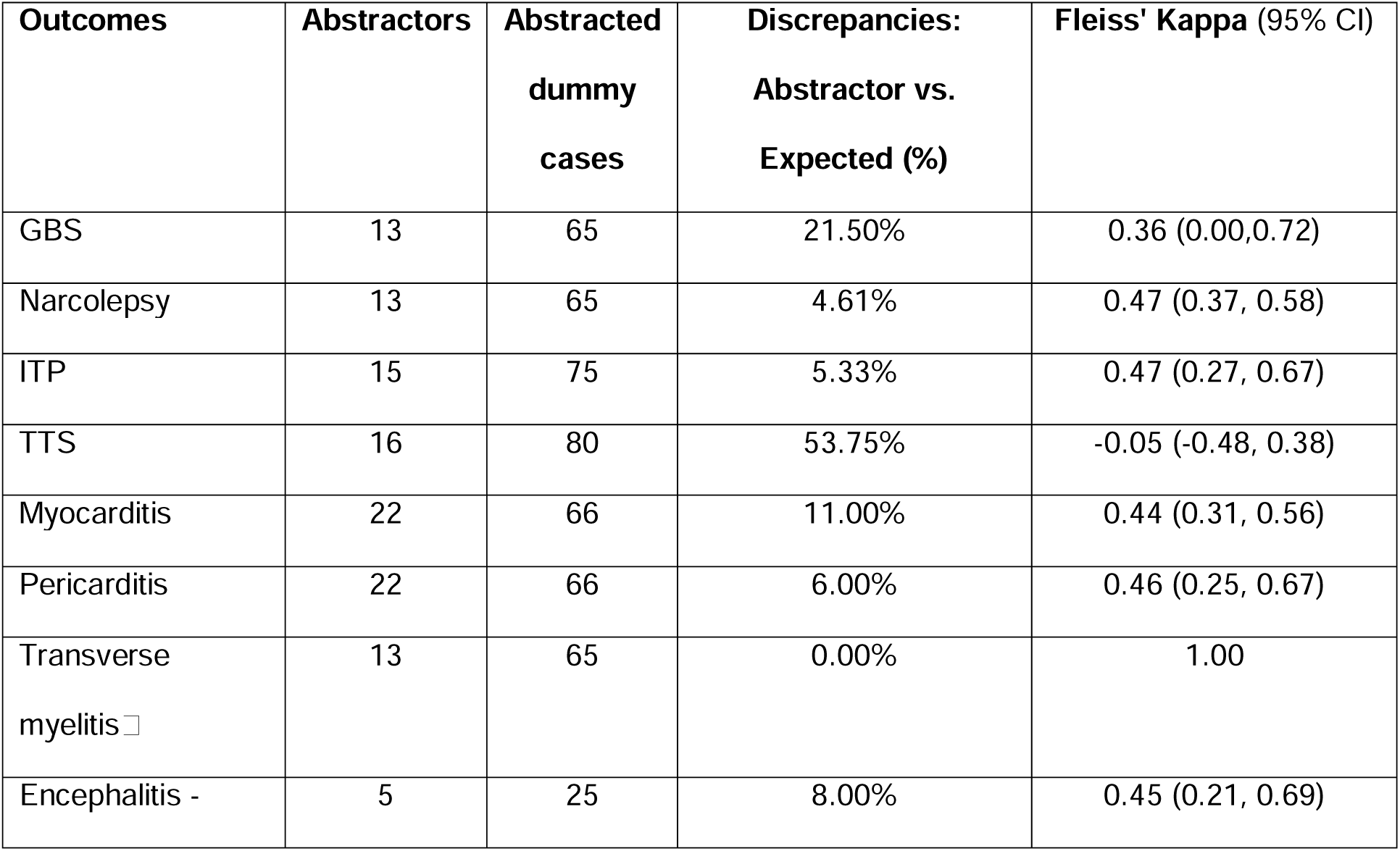
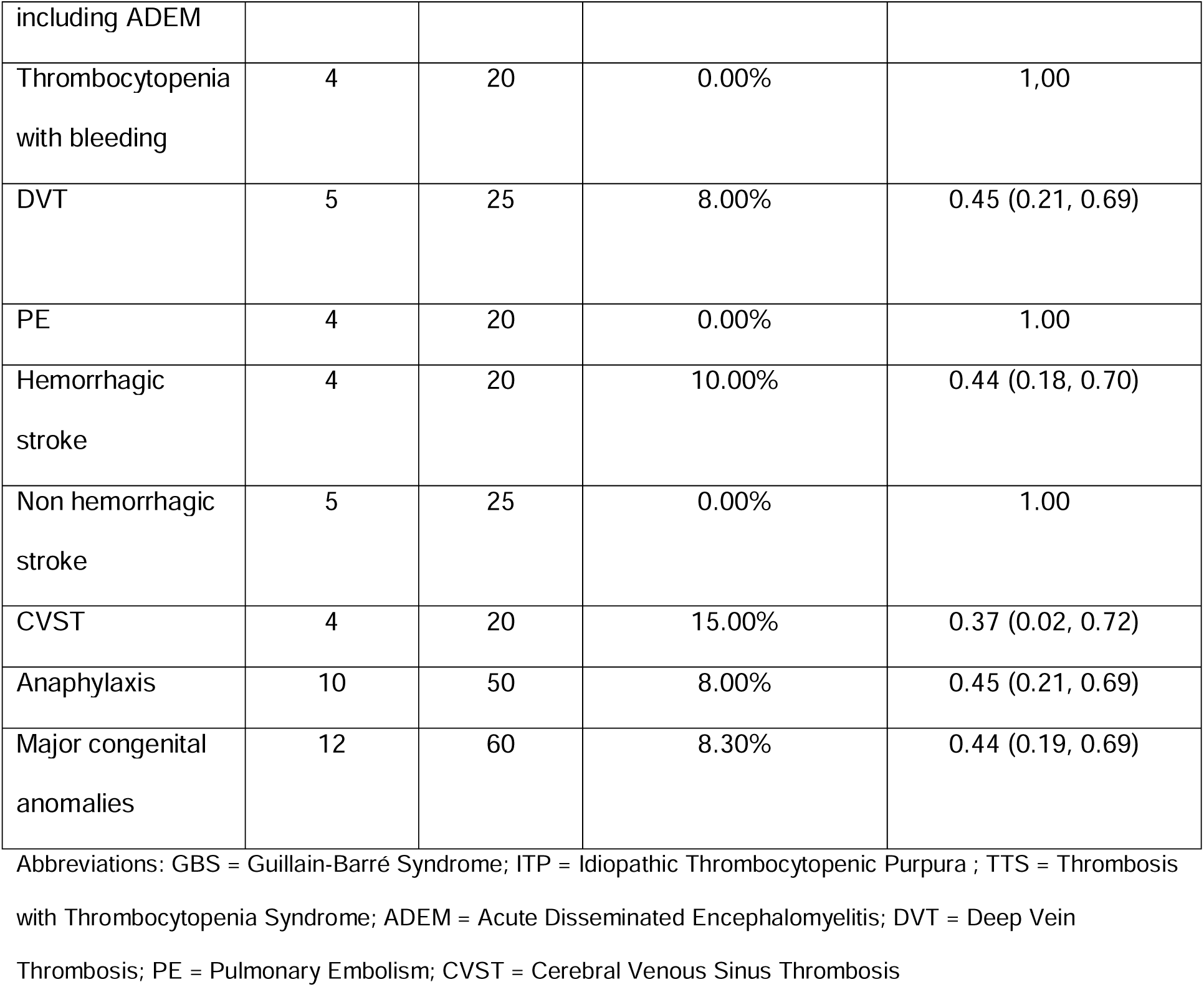
The distribution of abstractors across different outcomes with the distribution of discrepancies across abstracted dummy cases in the training.

The highest agreement among abstractors (Fleiss’ Kappa = 1.00) was observed for several outcomes, including transverse myelitis, thrombocytopenia with bleeding, PE, and non-hemorrhagic stroke.

In contrast, the highest number of discrepancies was found in the TTS cases, with 53.75% discrepancies and a Kappa of -0.05 (95% CI: -0.48 to 0.38), indicating no agreement. The GBS cases showed 21.50% discrepancies, with a Kappa of 0.36 (95% CI: 0.00 to 0.72), suggesting moderate agreement.

The remaining cases, such as narcolepsy, myocarditis, pericarditis, ITP, encephalitis including ADEM, hemorrhagic stroke, CVST, anaphylaxis, and major congenital anomalies, showed moderate to fair agreement.

For cases with significant discrepancies, such as in TTS, additional training meetings were scheduled to address the issues identified during dummy case abstraction. These sessions included a review of misclassified cases, clarification of complex criteria, for instance, interpretation of D-dimer values or alternative diagnoses, and discussion of recurring misunderstandings. Minor discrepancies, such as overlooking one of several required clinical symptoms or entering an incorrect event date due to a typographical error, were communicated via email with specific guidance to reduce errors. In total, three abstractors were removed from the validation process due to repeated errors and insufficient medical knowledge, which impacted their ability to meet validation standards.

Figure 2 illustrates the distribution of validation discrepancies across five major criteria categories: clinical symptoms, laboratory data, imaging evidence, other criteria, and alternative diagnosis, for each of the assessed outcomes. The most substantial number of discrepancies was observed in the laboratory data category, predominantly driven by TTS cases, where laboratory-related errors accounted for 17.60% of total discrepancies. These discrepancies were often due to inconsistent interpretation of laboratory values, for example, variation in D-dimer reporting units such as fibrinogen equivalent units (FEU) or milligrams per milliliter.

**Figure 2.**
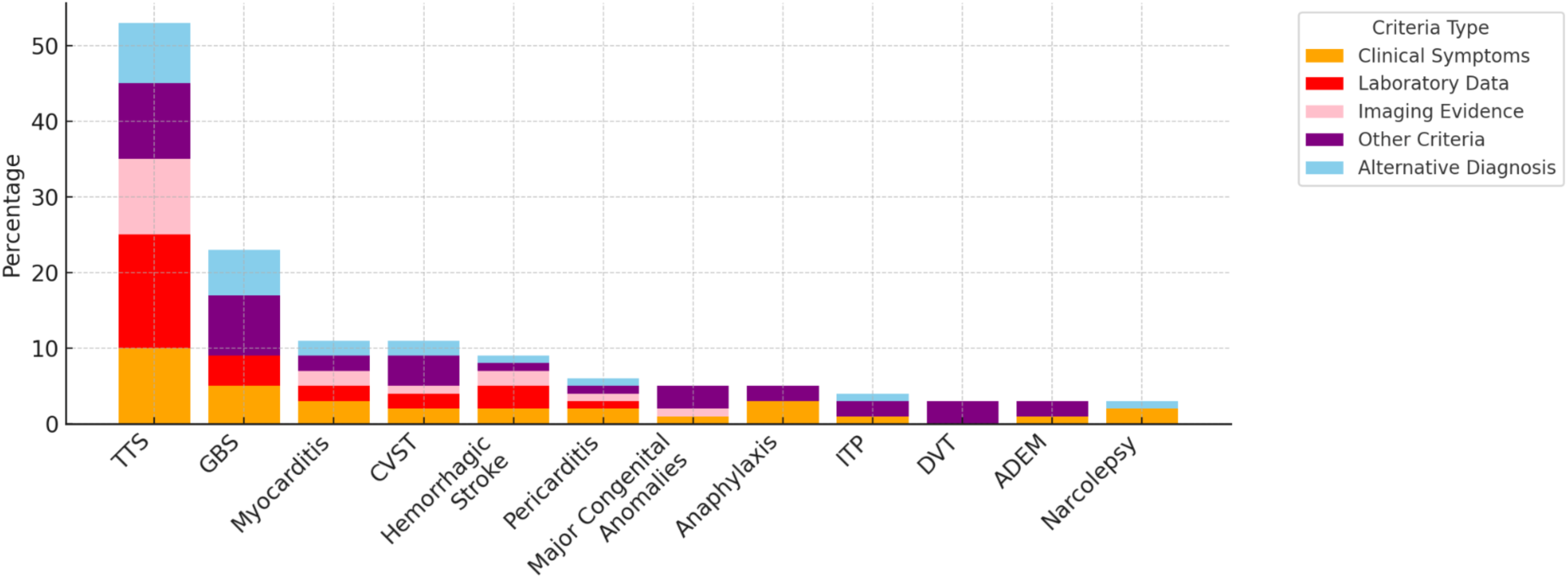
Distribution of validation discrepancies across criteria for different outcomes. TTS = Thrombosis with thrombocytopenia syndrome; GBS = Guillain-Barré syndrome; CVST = Cerebral venous sinus thrombosis; ITP = Idiopathic thrombocytopenic purpura; DVT = Deep vein thrombosis; ADEM = Acute disseminated encephalomyelitis

Errors in the alternative diagnosis and other criteria categories were more prominent in myocarditis, pericarditis, and GBS cases. Notably, myocarditis and pericarditis each had over five discrepancies attributed to alternative diagnoses. The GBS cases also had a high contribution from alternative diagnosis errors, totaling 12.00% of discrepancies for that event.

Discrepancies related to clinical symptoms were generally lower, but showed some variation, especially in TTS, Narcolepsy, and Anaphylaxis. For instance, Anaphylaxis showed 8.00% discrepancies due to errors in identifying clinical features. Additionally, Major Congenital Anomalies had 8.30% discrepancies, primarily attributed to misclassification under the “other criteria” category, such as incorrect assignment of the event date or misclassification of the anomaly type, including internal, external, or functional anomalies.

Overall, the distribution of discrepancies suggests that alternative diagnosis and other criteria introduce the greatest challenges for abstractors.

### Real Case Validation Outcomes

Following dummy-based training and reliability assessment, the pipeline was deployed to validate real potential cases across the five PASS studies. This resulted in the following confirmed case counts after abstraction and assignment of LOC: 153 cases of GBS, 83 cases of narcolepsy, 297 cases of ITP, and 243 cases of TTS were validated. Additionally, there were about 1570 cases of myocarditis, 2250 cases of pericarditis, and 41 cases of transverse myelitis. Other validated outcomes included 16 cases of acute disseminated encephalomyelitis including ADEM, 50 cases each of DVT and PE, 39 cases of hemorrhagic stroke, 50 cases of non-hemorrhagic stroke, 12 cases of CVST, 115 cases of thrombocytopenia with bleeding, 66 cases of anaphylaxis, and 237 cases of major congenital anomalies. Positive predictive values (PPVs) were calculated for these outcomes and used to adjust relative risk estimates in corresponding vaccine safety studies.

## Discussion

This study described how a harmonized validation process of outcomes was implemented in the VAC4EU validation pipeline. A key finding is that while BC case definitions and companion guides offer a standardized framework for vaccine safety assessment, they were originally developed for clinical or trial-based settings and often require adaptation when applied retrospectively to RWD. A major limitation is that BC definitions were highly clinical and rely on detailed physical examination findings, specific laboratory parameters, and imaging results, which are often unavailable, inconsistently recorded, or entirely absent in RWD sources. This structural mismatch affected abstractors’ ability to apply case definitions uniformly, especially when evaluating critical diagnostic elements such as laboratory values (e.g., variation in D-dimer units) or imaging findings. Additionally, it could be challenging. These findings are consistent with those of previous multinational validation studies, which have emphasized the inherent difficulty of applying trial-based or clinically detailed case definitions to observational studies relying on routinely collected health data ^31,32^. In particular, these studies highlighted that routinely collected data are often not structured to capture the full spectrum of clinical detail necessary to meet traditional regulatory-grade endpoint definitions. Even when adapted, such definitions may yield low PPVs due to this misalignment between clinical expectations and data reality. Although awareness of this mismatch is growing, it has yet to be fully reflected in routine validation practices in pharmacoepidemiology. This underscores the need for greater emphasis on developing fit-for-purpose definitions that are both clinically meaningful and feasible for use in RWD-based safety studies.

In addition to adapting existing BC definitions, the VAC4EU task force developed new definitions for outcomes not covered by BC, such as hemorrhagic stroke or thrombocytopenia with bleeding. These were created based on expert consensus and literature, and tailored to align with the structure and clinical content typically available in real-world data sources. A key challenge was balancing clinical specificity with feasibility across heterogeneous databases. While these new definitions were not externally validated, they were refined during implementation based on feedback from abstractors. This process highlights the importance of designing fit-for-purpose definitions as a core component of any scalable validation framework.

The Vaccine Safety Datalink (VSD) also highlights the need for thorough clinical review to ensure the accuracy of diagnoses, especially in cases where automated data may not fully capture complex clinical histories ^33^. This also suggests that targeted adjustments in training materials and clarification within specific validation criteria could improve further case validation. The Canadian Network for Observational Drug Effect Studies (CNODES) similarly emphasizes the role of clinical expertise in navigating complex data sets and maintaining the integrity of observational studies ^34^. Additionally, the distribution of errors highlights the importance of maintaining detailed guidance on laboratory data and alternative diagnosis criteria, especially for complex or multi-faceted outcomes like TTS, GBS, and Myocarditis. Fleiss’ Kappa calculations further underscore the challenges faced in dummy cases validation. Specifically, the TTS had the highest level of disagreement, with a low Kappa value, indicating poor agreement between abstractors. This aligns with the earlier finding that decisions around alternative diagnoses, such as chronic thrombocytopenia, were difficult to assess. The GBS cases also demonstrated moderate agreement with a Kappa value suggesting that inconsistencies in interpretation were more frequent in cases with ambiguous or complex diagnostic criteria. On the other hand, several outcomes, such as transverse myelitis, thrombocytopenia with bleeding, PE, and non-hemorrhagic stroke, showed full agreement among abstractors.

The multinational structure of the VAC4EU network introduced further complexity. All training materials and forms were developed in English to support broad accessibility across language settings, though minor interpretation differences may have arisen for non-native speakers.

Additionally, the availability of diagnostics, treatment practices, and coding systems varied across countries and data sources, occasionally limiting the abstractors’ ability to complete certain fields in the eDCFs. This variation highlights the importance of designing abstraction tools with sufficient flexibility and aligning questions with the typical data content of participating databases.

These findings highlight the need for targeted training and clarification of diagnostic criteria, particularly for outcomes where abstractor decisions were more prone to variability. The moderate to poor agreement observed in some outcomes reflects the challenges in achieving harmonized results in situations where clinical judgment and alternative diagnoses play a significant role. Fleiss’ Kappa values serve as an important tool in pinpointing areas where additional guidance or clarification is needed to ensure consistency across abstractors.

Second, considerable effort was put into creating REDCap forms and verification of the logic. Making such forms and logic available in a digitized format—whether through Brighton Collaboration or coordinated networks, could facilitate implementation across networks, reduce duplication of effort, and improve consistency in applying the definitions. Moreover, incorporating insights from field implementation, including training feedback and validation challenges, would enhance the practical applicability of these definitions in RWD settings. Therefore, VAC4EU served as a “living lab” for the safety platform for emergency vaccines (SPEAC) project, offering feedback and applied experience to help refine future tools and guidance from the Brighton Collaboration. Third, deployment in studies was possible in a harmonized and distributed manner. The VAC4EU validation pipeline provided support and quality control during the implementation of case validation phase in safety studies.

Meetings with abstractors provided opportunities to address their challenges and maintain consistency across DEAPs. This close communication and feedback mechanism was essential for managing the complexities of validating diverse outcomes. These efforts ensured that distributed abstractors were supported in navigating complex scenarios and aligned with the study’s standards.

The VAC4EU validation pipeline offers a scalable and adaptable model for future studies. By identifying challenges, emphasizing the critical role of medical expertise, and ongoing support through structured feedback, VAC4EU has set a standard for implementation of validation according to Brighton Collaboration criteria in retrospective use of RWD in vaccine safety monitoring. This process was designed to counter the inherent challenges of RWD, including heterogeneity across datasets, varying levels of data completeness, and the need for consistent application of standardized definitions. The pipeline incorporates a series of interdependent steps, each aimed at promoting data integrity, enhancing abstraction accuracy, and ensuring harmonized validation across multiple DEAPs.

### Strengths and Limitations

A major strength of this study is the application of a harmonized and scalable validation framework across a large, multinational network using RWD. The use of standardized training materials, structured data collection tools, and systematic adjudication processes ensured consistency and comparability across diverse settings. The development of novel case definitions in the absence of Brighton Collaboration references also demonstrated the task force’s adaptability to RWD constraints. However, several limitations should be noted. The dummy cases used for training were narrative-based and may not fully reflect the structure of actual data extracts encountered during study implementation. Furthermore, differences in data availability, language, and healthcare practices across DEAPs may have affected abstraction consistency.

## Conclusion

In conclusion, The VAC4EU validation pipeline establishes a standardized and scalable framework for validating vaccine safety outcomes using RWD. By adapting BC case definitions to the realities and constraints of RWD, and by implementing comprehensive abstractor training with ongoing feedback, this approach enhances the accuracy, consistency, and reproducibility of outcome adjudication across multiple sites and countries. This harmonized validation process supports robust post-marketing vaccine safety monitoring and serves as a replicable model for future multinational collaborative studies leveraging heterogeneous RWD sources.

## Data Availability

All data produced in the present study are available upon reasonable request to the authors

## Authors’ Contributions

MS conceptualized the study and supervised all stages of the validation pipeline. AD developed the manuscript, conducted the case validation pipeline, and coordinated co-author feedback. JF, CR, AA, FA, and DW contributed to the case validation task force by developing materials and supporting the implementation of the pipeline. TL, JVB, VE, JW, TA, LP, AG, LS, LZ, MS, AAD, BP, MZ, JC, FV, and KM coordinated the data abstraction process and contributed to quality control. DW and FA advised on methodology and critically revised the manuscript. All authors reviewed and approved the final version of the manuscript and agree to be accountable for all aspects of the work.

## Acknowledgments

The authors would like to thank all members of the VAC4EU network who contributed to the development, testing, and implementation of the validation pipeline. We are especially grateful to the members of the validation task force for their input on adapting case definitions, creating training materials, and coordinating abstraction activities across participating data sources.

We would also like to thank the following abstractors for their invaluable contributions to the manual validation process across the various outcomes and data sources included in this study: Rachel Aakerøy, Ashi Sarfraz Ahmad, Olaug Marie Bruheim Reiakvam, Juanjo Carreras, Costanza Di Chiara, María Díaz López, Edison Daniel Valenzuela Cumba, Sandeep Dhanda, Miranda Davies, Pablo Daniel Estrella Porter, Sílvia Fernández García, María Luisa Gil Canela, María Gine, Nils Erik Gilhus, Anyuli Gracia Gutiérrez, Eva Jara Castillejo, Christopher Steph Inchley, Laurits Juhl Heinsen, Parinaz Heydari, Thom Lysen, Aida Moreno Juste, Kathryn Morton, Marta Pastor Sanz, Sergio Pascual Viciedo Mata, Roberto Peribañez García, Lina Pérez Breva, Oscar Oelrich Rosenkrantz, Martín Solórzano, Esther Soriano García, Victor Hejgaard Sørensen, Christian Thaulow, Jesse Van den Berg, and Jannik Wheler.

Their careful review and commitment to consistency played a key role in achieving the quality and reproducibility of the validation process.

This study is based in part on data from the Clinical Practice Research Datalink (CPRD) (study protocol numbers: 21_000714, 21_000535, 23_003471) obtained under licence from the UK Medicines and Healthcare products Regulatory Agency. The data is provided by patients and collected by the NHS as part of their care and support. The interpretation and conclusions contained in this study are those of the author(s) alone.

